# Correlation of Social Determinants of Health and Chronic Nonspecific Low Back Pain; A Brief Report

**DOI:** 10.1101/2023.12.06.23299619

**Authors:** Alireza Ebrahimi, Hyunjoon Rhee, Atta Taseh, Hye Chang Rhim, Soheil Ashkani-Esfahani

## Abstract

**Background:** Low back pain (LBP) ranks as one of the top contributors to disability-adjusted life years worldwide. Social determinants of health (SDoH) encompass a range of factors that play a pivotal role in shaping healthcare outcomes. We aimed to examine the differences in SDoH between patients with acute and chronic non-specific low back pain (NSLBP).

**Methods:** This study was a retrospective case-control study on patients who were admitted to four tertiary hospitals affiliated with Massachusetts General Hospital. Relevant ICD-9 and ICD-10 codes were used to detect patients with LBP. Patients with specific diagnoses such as degenerative joint diseases or primary or secondary bone tumors were excluded. SDoH variables along with the Social Vulnerability Index (SVI) and Area Deprivation Index (ADI) were collected. Wilcoxon rank sum and Chi-square tests were performed to assess the difference between the cohorts.

**Results:** Of 2,278 patients who were labeled as chronic LBP, 17 were included. The number of female patients was significantly higher (P < 0.05). Furthermore, we didn’t observe any considerable difference between these two groups of patients regarding the SDoH, SVI, and ADI values.

**Conclusion:** The prevalence of chronic NSLBP in tertiary healthcare centers might be lower. SDoH, ADI, and SVI seem not to contribute to the chronicity of NSCLBP. Future research is needed to further our understanding of the risk factors associated with NSLBP, particularly in different healthcare settings.

**Significance/Implications:** Although, SDoH variables are mentioned as risk factors, considering the clinical setting when applying the knowledge seem to play an important role.

## Introduction

Low back pain (LBP) is among the world’s leading causes of disability-adjusted life years (DALYs) (GBD 2019 Diseases and Injuries Collaborators, 2020). Nearly 4 out of 5 adults in the United States experience LBP at least once in their lifetime (Urits et al., 2019). The estimated cost of treating the morbidities resulting from LBP exceeds 600 billion annually (Becker & Childress, 2019). Although LBP is usually self-limited, about 30% becomes chronic lasting more than 12 weeks (Patrick et al., 2016). It is worth mentioning that the majority of the economic and psychosomatic burden is due to its chronicity (Patrick et al., 2016). Patients with no established underlying cause for LBP, such as fractures, infections, and tumors, are labeled non-specific LBP (NSLBP) with a prevalence of ∼5% among all LBP patients (Karran et al., 2020). The physicians might cautiously diagnose the patients with chronic NSLBP when ruling out all serious causes of LBP.

Early intervention was found to improve patient-reported pain scores and functional outcomes in NSLBP (Yeganeh et al., 2017). Previous studies have recognized different risk factors such as obesity and smoking to be associated with the chronicity of LBP (Shemory et al., 2016; Stevans et al., 2021). According to the literature, the majority of symptoms in specific LBP are rooted in underlying medical or pathoanatomical reasons, whereas the symptoms of NSLBP were found to be highly related to psychosomatic problems and individual characteristics of the patients (Kent & Kjaer, 2012; Werneke & Hart, 2001).

Social determinants of health (SDoH) refer to the social, economic, educational, cultural, and environmental factors that could affect individuals’ health (WHO, 2023). These factors contribute to healthcare outcomes and could lead to health inequities, which are defined as preventable or modifiable differences in the health status of various subgroups of populations caused by socioeconomic status (SES) (WHO, 2023). The Global Commission on Social Determinants of Health (CSDH) has highlighted the importance of tackling inequities by improving individuals’ living conditions, addressing inequitable resources, and understanding and measuring the problem (WHO, n.d.). During the past few decades, the impacts of SDoH on various musculoskeletal disorders, specifically LBP, have been studied (Guillemin et al., 2014). A recent systematic review of multiple studies reported that educational and economic status had a positive correlation with spine surgery outcomes in patients with LBP (Yap et al., 2022).

Another review with a much larger population mentioned that a range of interdependent and independent correlations could be found between LBP management outcomes and SDoH variables, including gender, marital status, and place of living (Karran et al., 2020). Previous studies showed that social determinants of health might define the outcomes of patients with chronic LBP, however, the relationship of these factors to chronic NSLBP is yet to be elaborated. Based on these pieces of evidence, and to the best of our knowledge, studies on the correlation of SDoH and the outcomes of NSLBP are scarce. We assumed that these patients, since the underlying cause for their LBP has not been well defined, are at higher risk of being neglected, mismanaged, and thus, facing further complications. Hence, understanding the factors that could influence the management process of these patients seems to be of great importance. The current study aimed to compare SDoH in patients with acute and chronic NSLBP. We hypothesized that SDoH could play a predictive role in chronicity in NSLBP and help providers plan for prevention or early intervention.

## Material and Methods

### 1.1. Study Design and Population

This study was a retrospective case-control study on patients who were admitted to four tertiary hospitals and clinics affiliated with our institution between the years 2016 and 2022. We used the relevant ICD-9 and ICD-10 codes to detect patients with LBP in our enterprise data warehouse (EDW) using the Research Patient Data Registry Application (RPDR) (Supplementary 1). We extracted all the encounters of patients who were 18 years old or older and had complaints of LBP in their orthopaedic or physical therapy (PT) visits. The patients with specific diagnoses such as degenerative joint diseases, disk fractures, scoliosis, disk stenosis, and primary or secondary bone tumors affecting disks were excluded. We also excluded the patients who didn’t have at least two visits and the patients with a history of spine surgery. The inclusion and exclusion criteria are summarized in Table 1. Finally, the resulting dataset was reevaluated manually by two expert orthopaedic researchers.

**Table 1.**
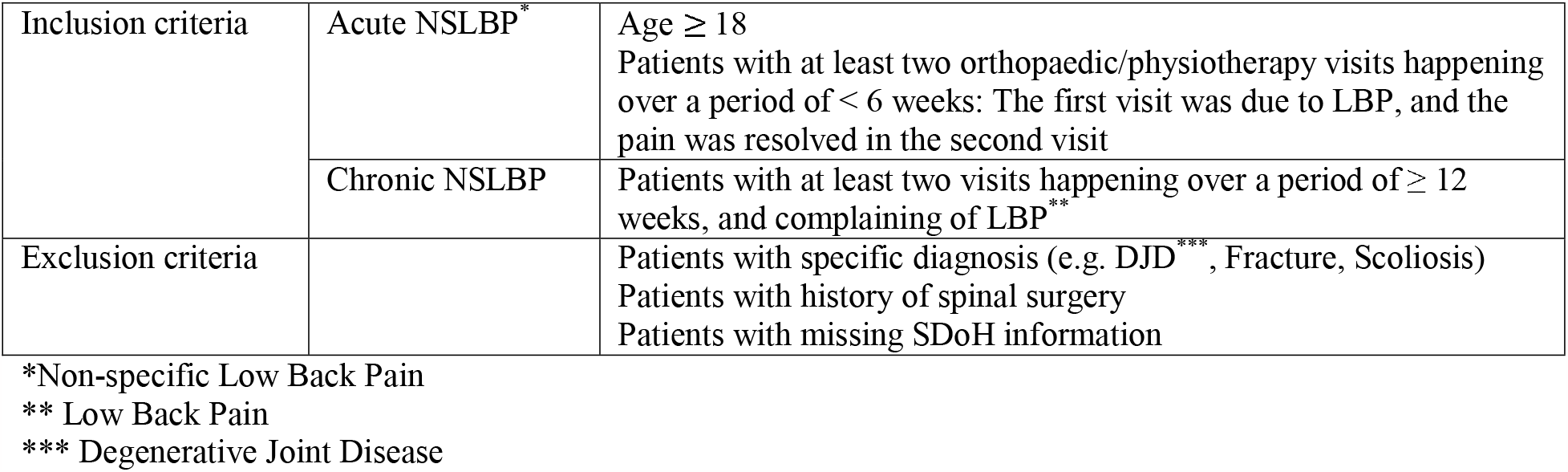
Inclusion and exclusion criteria.

### 1.2. Social Determinants of Health

We gathered SDoH variables including age, gender, marital status, race, ethnicity, type of insurance, and variables related to the lifestyle and daily habits of the patients. Social vulnerability index (SVI) and area deprivation index (ADI) are based on the Zip code of patients’ location and were collected from open-source databases (Angelidou et al., 2021).

The social vulnerability index (SVI), which is a composite of different factors affecting the outcomes of patients with different diseases, was collected from the Agency for Toxic Substances and Disease Registry (ATSDR) website (ATSDR, 2022). SVI encompasses 4 main categories including, socioeconomic status (SES), household composition and disability, minority status and language, and housing and transportation (16). Patients’ scores for SES, household characteristics, minority status, and transportation type were also gathered from the same resource (16). It is noteworthy to mention that each of these categories also contains different factors. Subsequently, SVI evaluates 15 criteria based on their relative influence (weights) on health. The metric scores the individuals from 0 (defined as lowest vulnerability) to 1 (defined as highest vulnerability) based on ZIP code Tabulation Areas (ZCTAs) (16). Additionally, the national percentile and state decile area deprivation index (ADI) of the patients were gathered from the online resource designed by the Center for Health Disparities Research, University of Wisconsin (University of Wisconsin, n.d.). ADI values are provided as state decile rankings at a block group level from 1 to 10, where 1 being the lowest level of disadvantage and 10 being the highest level of disadvantage. Furthermore, national percentiles of ADI scores were collected, where the lowest score of 1 shows the least disadvantaged group, and the highest score of 100 shows the most disadvantaged group. These pieces of data are based on counties, which is more general than the geographical specificity of SVI, and are calculated according to the American Community Survey (ACS) 2020 (17). Although SVI and ADI are more comprehensive and are becoming more popular than crude data of median income, we also gathered the patients’ median income based on Zip code from the United States Census Bureau website (United States Census Bureau, n.d.).

### 1.3. Statistical Analysis

The descriptive measures were reported as numbers, percentages, median, and interquartile range (IQR) where applicable. Wilcoxon rank sum and Chi-square tests were performed to assess the difference between the cohorts. All statistical analyses were done using the SPSS® Software version. 28, and P-value < 0.05 was considered statistically significant.

### 1.4. Ethical consideration

All the patients’ identifiers were removed in this study, and the protocol of the study was approved by the institutional review board (IRB no. 2023P000728).

## Results

Overall, 51,636 patients were diagnosed with LBP between 2012 and 2022 at four tertiary hospitals affiliated with our institution. Of 2,278 patients who were labeled as chronic LBP using the aforementioned methodology, 17 patients were chronic NSLBP. From those with acute NSLBP (n = 1115), a randomly selected cohort of 17 patients was used as a comparison group (Figure 1).

**Figure 1.**
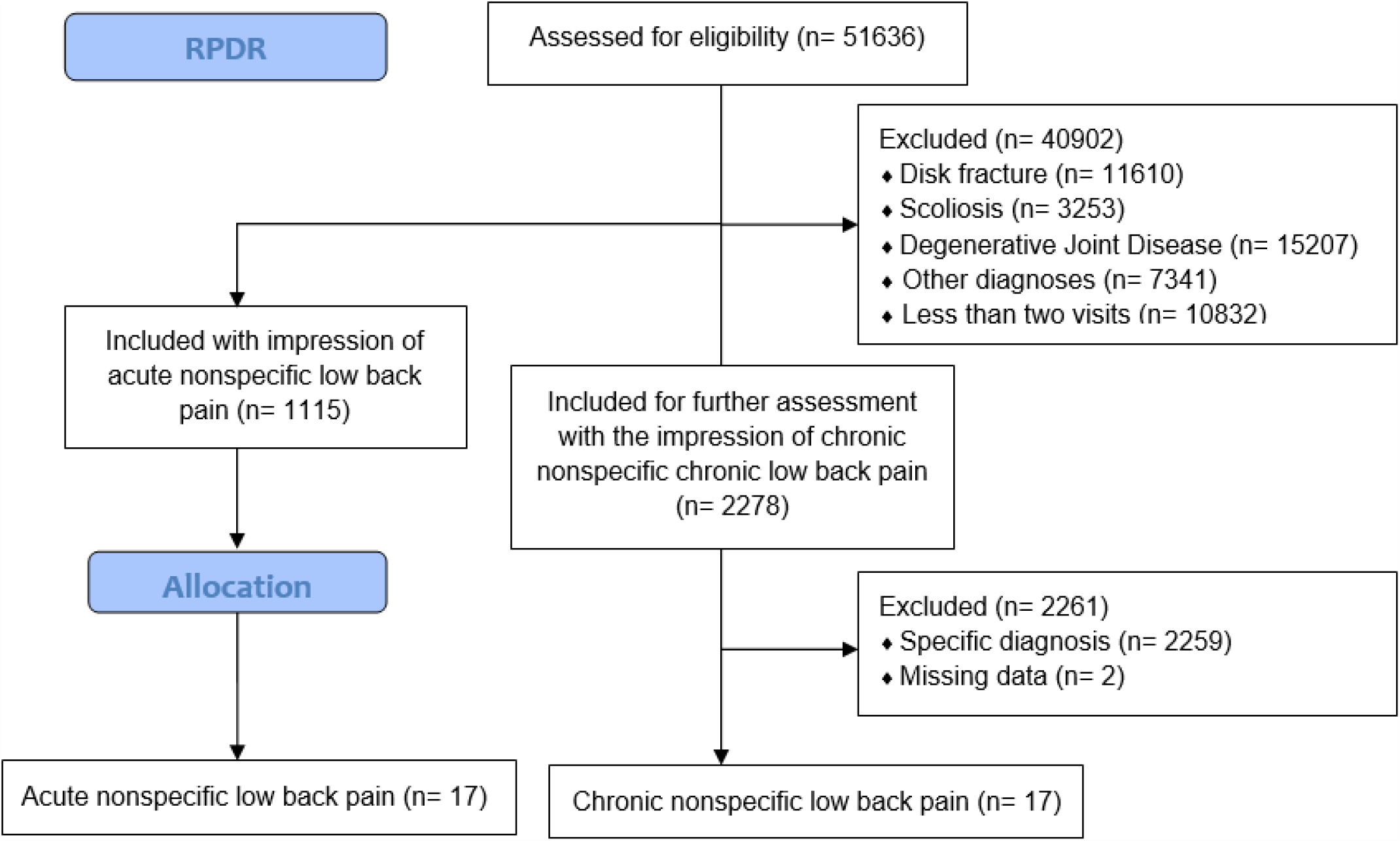
Recruitment Flowchart

Table 2 represents the demographics of patients who had acute and chronic NSLBP. The results of the present study showed that the number of female patients with chronic NSLBP was significantly higher than their male counterparts (P < 0.05). Furthermore, we didn’t observe any considerable difference between these two groups of patients in smoking and alcohol use, SVI, SES, household characteristics, minority status, housing type and transportation, ADI, and median income (Table 3).

**Table 2.** Comparison of the demographic data of patients with acute versus chronic non-specific low back pain.

| Variable | Chronic (n = 17) | Acute (n = 17) | P-value * |
| --- | --- | --- | --- |
| Age (years old) | Median (Q1-Q3)<br>61 (45.5-68.5) | Median (Q1-Q3)<br>52 (45.5-62.5) | 0.538 |
| Gender | N (Percentage)<br>Male: 6 (35.3%)<br>Female: 11 (64.7%) | N (Percentage)<br>Male: 12 (70.6%)<br>Female: 5 (29.4%) | 0.039 |
| Marital status | N (Percentage)<br>Single: 4 (23.5%)<br>Married: 8 (47.1%)<br>Widowed/Divorced: 5 (29.4%) | N (Percentage)<br>Single: 10 (58.8%)<br>Married: 5 (29.4%)<br>Widowed/Divorced: 2 (11.8%) | 0.445 |
| Race | N (Percentage)<br>White: 12 (70.6%)<br>Black: 1 (5.9%)<br>Asian: 1 (5.9%)<br>Other: 3 (17.6%) | N (Percentage)<br>White: 13 (76.5%)<br>Black: 0<br>Asian: 4 (23.5%)<br>Other: 0 | 0.12 |
| Ethnicity | N (Percentage)<br>Not Hispanic: 15 (88.2%)<br>Hispanic: 1 (5.9%)<br>Other: 1 (5.9%) | N (Percentage)<br>Not Hispanic: 16 (94.1%)<br>Hispanic: 0<br>Other: 1 (5.9%) | 0.597 |
| Insurance | N (Percentage)<br>Medicare: 6 (35.3%)<br>Private: 10 (58.8%)<br>Other: 1 (5.9%) | N (Percentage)<br>Medicare: 6 (35.3%)<br>Private: 11 (64.7%)<br>Other: 0 | 0.592 |
| BMI | Median (IQR)<br>26.3 (20.42-29.13) | Median (IQR)<br>23.9 (20.82-28.91) | 0.657 |
| Smoking | N (Percentage)<br>Never: 10 (58.8%)<br>Former: 5 (29.4%)<br>Smoker: 2 (11.7%) | N (Percentage)<br>Never: 14 (82.3%)<br>Former: 2 (11.7%)<br>Smoker: 1 (5.9%) | 0.319 |
| Alcohol | N (Percentage)<br>Never: 7 (41.1%)<br>Occasionally: 1 (5.9%)<br>Regularly: 9 (52.9%) | N (Percentage)<br>Never: 11 (64.7%)<br>Occasionally: 2 (11.7%)<br>Regularly: 4 (23.5%) | 0.207 |

\* Wilcoxon rank sum test was performed for the comparison of numerical variables, and Chi-square test were conducted for nominal variables.

**Table 3.** Social determinants of health in patients with acute and chronic non-specific low back pain.

| **Variable** | **Chronic**<br>**Median (Q1-Q3)** | **Acute**<br>**Median (Q1-Q3)** | **P-value**<br>**\*** |
| --- | --- | --- | --- |
| Overall Social Vulnerability Index (SVI) ** | 0.28 (0.27-0.78) | 0.28 (0.26-0.58) | 0.379 |
| Socioeconomic Status ** | 0.19 (0.12-0.65) | 0.19 (0.11-0.44) | 0.53 |
| Household Characteristics ** | 0.22 (0.11-0.35) | 0.22 (0.06-0.35) | 0.851 |
| Minority Status ** | 0.63 (0.58-0.84) | 0.63 (0.55-0.66) | 0.753 |
| Housing Type and Transportation ** | 0.75 (0.70-0.94) | 0.75 (0.53-0.82) | 0.615 |
| Area Deprivation Index (National Percentile) *** | 23 (11.5-30) | 23 (9-45) | 0.381 |
| Area Deprivation Index (State Decile) *** | 6 (3-7) | 6 (2-8.5) | 0.588 |
| Median Income (IQR) **** | 106153 (69276-116865) | 90315 (68230-115749) | 0.653 |

\* Wilcoxon rank sum test was performed for the comparison.
\*\* The data relating to these variables were gathered from Agency for Toxic Substances and Disease Registry (ATSDR) website: https://www.atsdr.cdc.gov/placeandhealth/svi/interactive_map.html
\*\*\* The data relating to these variables were gathered from Center for Health Disparities Research, University of Wisconsin website: https://www.neighborhoodatlas.medicine.wisc.edu/
\*\*\*\* The data for these variables were gathered from the United States Census Bureau website: https://data.census.gov/

## Discussion

The pathophysiology of NSLBP and the risk factors contributing to the development and progression of the disease are still unclear. There is also a lack of data regarding the epidemiology of NSLBP among both acute and chronic cases. A few studies have delved into assessing the prevalence of chronic NSLBP and reported a range of 7-80% (Chiwaridzo & Naidoo, 2014). In the present study, the ratio of patients with chronic NSLBP to all the patients found to have chronic LBP was about 1%. One possible explanation for the lower rate could be that our centers are tertiary facilities, and there is a higher possibility that patients are diagnosed with an actual diagnosis rather than NSLBP. Among different SDoH-related factors, gender showed a significant difference between the acute and chronic cohorts of NSLBP. While our low population can limit the generalizability of our outcomes, our results showed that the female gender can be a predictor of chronicity in our population with NSLBP. Consistent with our outcomes, Bento et al. showed that the prevalence of LBP is considerably higher in female (60.9%) than the male (39.0%) participants of their cross-sectional study (Bento et al., 2020). Lu et al. investigated multiple risk factors and reported female gender along with ten other factors that are of predictive value (Lu et al., 2023). Another study by Gok et al. also confirmed that gender has an influence on inflammatory criteria of LBP (Gok et al., 2022).

Previous studies on the effect of SDoH on LBP are mostly focused on a specific domain and it is difficult to draw conclusions from them because of the controversial results. Several studies underlined the effect of educational level and income status in this patient population (Park et al., 2023; Takahashi et al., 2018). A recent systematic review and meta-analysis of the effect of SDoH on LBP by Karran et al. showed that disparities in socioeconomic status and educational attainment significantly contribute to chronic LBP (5). On the contrary, Ramond et al included 18 cohorts in a systematic review and reported no association between most social and socio-occupational factors with chronic low back pain (Ramond et al., 2011). Our results also did not reveal any significance regarding the effect of SVI and its subscales including socioeconomic status on the chronicity of the condition. One possibility could be that because our data were collected in a referral setting, there might exist a disparity in referring patients to such centers. According to a study by Olah et al, the likelihood of being offered a primary care appointment is significantly higher when patients represent themselves as having high socioeconomic status (Olah et al., 2013).

This study has several limitations to be mentioned. Despite the advantages of SVI and ADI, accurately predicting the score for each individual was impossible as the data gathered according to ZCTAs. The individuals might live in a neighborhood that is not representative of their income, education, and other determinants of health. Also, SVI scores are given to geographical locations that are grouped into counties, which in many cases cover a vast region that may elicit disparities of the social status within the counties. Our study was done on a limited cohort that might not represent different populations at national and global levels. The centers involved in the current study are considered referral centers, and the rate of patients with NSLBP might not be representative of the patients receiving care in primary care centers. Further studies on larger sample sizes are suggested to address these limitations.

## Conclusion

In conclusion, the risk factors contributing to the chronicity of NSLBP are still to be investigated, and there is still a lack of comprehensive data on its epidemiology. The present study suggests that the prevalence of chronic NSLBP in tertiary healthcare centers might be lower compared to primary and secondary clinics. Despite the limitations of this low sample size study, our findings indicate that female gender could be considered as a risk factor for chronicity of NSLBP patients. Further research is needed to further our understand of the risk factors associated with NSLBP, particularly in different patient populations and healthcare settings.

## Data Availability

All data produced in the present study are available upon reasonable request to the authors

## Research funding

None declared.

## Author contributions

All authors have accepted responsibility for the entire content of this manuscript and approved its submission.

## Informed consent

Informed consent was obtained from all individuals included in this study.

## Ethical approval

The local Institutional Review Board has approved the protocol of the study (IRB no. 2023P000728).

