## Supplemental Table 1 for "Correlation of Social Determinants of Health and Chronic Nonspecific Low Back Pain; A Brief Report"

**Supplementary**

**Suppl 1. ICD-9/ICD-10/CPT codes utilized to detect the patients with LBP**

| **ICD-9** | | **ICD-10** | | **CPT** | |
| --- | --- | --- | --- | --- | --- |
| **Code** | **Definition** | **Code** | **Definition** | **Code** | **Definition** |
| S30.0 | Contusion back | M54.5- | Low back pain, unspecified | 74000 | Abdomen 1-view |
| 724.2 | Low back pain, unspecified | S39.012 | Low back strain | 74020 | Abdomen 2- view |
| 846.0 | Lumbosacral strain | M54.51 | Vertebrogenic low back pain | 74022 | Abdomen 3- view |
| 847.2 | Low back strain | M54.59 | Low Back Pain, Site Unspecified | 72120 | L-spine flexion and extension minimum 4 |
| 722.1 | Lumbago due to intervertebral disc displacement | M51.2 | Lumbago due to intervertebral disc displacement | 72110 | L-spine minimum 4 views |
| 724.8 | Muscle spasm back | M54.4- | Lumbago with sciatica | 72020 | L-spine 1 view |
| 724.5 | Chronic low back pain | G89.29 | Chronic low back pain | 72100 | L-spine 2 or 3 views |
| 729.1 | Musculoskeletal back pain | G59.29 | Chronic intractable low back pain | 72010 | Spine complete 2 views |
| 922.3 | Contusion back | M54.9 | Dorsalgia Spine, unspecified | 72010 | x-ray spine entire |
| 722.73 | Intervertebral disc disorders with myelopathy, lumbar region | G89.21 | Chronic Pain in spine due to Trauma | 72020 | x-ray spine, 1 view |
| 722.52 | Other intervertebral disc degeneration, lumbar region | M51.06 | Intervertebral disc disorders with myelopathy, lumbar region | 72040 | xray spine cervical 2-3 views |
| 722.93 | Intervertebral disc disorders with radiculopathy, lumbar region | M51.07 | Intervertebral disc disorders with myelopathy, lumbosacral region | 72080 | x-ray spine thoracolumbar 2 views |
| 722.93 | Other lumbar intervertebral disc disorders | M51.36 | Other intervertebral disc degeneration, lumbar region | 72090 | x-ray spine thoracolumbar supine and standing |
| 722.93 | Unspecified lumbar intervertebral disc disorder | M51.37 | Other intervertebral disc degeneration, lumbosacral region | 72100 | x-ray spine lumbosacral 2-3 views |
| 724.02 | Intervertebral disc stenosis of neural canal of lumbar region | M51.16 | Intervertebral disc disorders with radiculopathy, lumbar region | 72110 | x-ray spine lumbosacral 4+ views |
| 724.02 | Osseous and subluxation stenosis of intervertebral foramina of lumbar region | M51.17 | Intervertebral disc disorders with radiculopathy, lumbosacral region | 72114 | x-ray spine lumbosacral complete |
| 721.9 | Spondylosis, unspecified, without mention of myelopathy | M51.86 | Other lumbar intervertebral disc disorders | 72120 | x-ray spine lumbosacral bending only |
| 721.41 | Other spondylosis with myelopathy, thoracolumbar region | M51.87 | Other lumbosacral intervertebral disc disorders |  |  |
| 721.41 | Other spondylosis with radiculopathy, thoracolumbar region | M51.96 | Unspecified lumbar intervertebral disc disorder |  |  |
| 721.3 | Other spondylosis with radiculopathy, lumbosacral region | M51.97 | Unspecified lumbosacral intervertebral disc disorder |  |  |
| 721.2 | Spondylosis without myelopathy or radiculopathy, thoracolumbar region | M99.53 | Intervertebral disc stenosis of neural canal of lumbar region |  |  |
| 722.5 | Degeneration thorc/lumbar disc | M99.54 | Intervertebral disc stenosis of neural canal of lumbosacral region |  |  |
| 722.52 | Degeneration, lumbar/lmbsac disc | M99.63 | Osseous and subluxation stenosis of intervertebral foramina of lumbar region |  |  |
| 737 | Kyphoscoliosis and scoliosis | M99.73 | Connective tissue and disc stenosis of intervertebral foramina of lumbar region |  |  |
| 805 | Fracture of vertebral column without mention of spinal cord lesion | M48.06 | Spinal stenosis, lumbar region |  |  |
| 806 | Fracture of vertebral column with spinal cord lesion | M48.05 | Spinal stenosis, thoracolumbar region |  |  |
| 170.9 | Primary malignant neoplasm of bone and articular cartilage: vertebrae | M48.07 | Spinal stenosis, lumbosacral region |  |  |
| 213.2 | Benign neoplasm of bone and articular cartilage: Spine | M47.9 | Spondylosis, unspecified, without mention of myelopathy |  |  |
| 733.00 | Age-related osteoporosis without current pathological fracture | M47.1 | Other spondylosis with myelopathy |  |  |
| 732.8 | Adult osteochondrosis of spine, site unspecified | M47.15 | Other spondylosis with myelopathy, thoracolumbar region |  |  |
| 015.00 | Tuberculosis of spine | M47.16 | Other spondylosis with myelopathy, lumbar region |  |  |
| 721.5 | Kissing spine, site unspecified | M47.25 | Other spondylosis with radiculopathy, thoracolumbar region |  |  |
| 756.15 | Fusion of spine, site unspecified | M47.26 | Other spondylosis with radiculopathy, lumbar region |  |  |
| 721.6 | Ankylosing hyperostosis [Forestier], site unspecified | M47.27 | Other spondylosis with radiculopathy, lumbosacral region |  |  |
| 720.81 | Other infective spondylopathies, site unspecified | M47.815 | Spondylosis without myelopathy or radiculopathy, thoracolumbar region |  |  |
| 720.9 | Unspecified inflammatory spondylopathy, site unspecified | M47.816 | Spondylosis without myelopathy or radiculopathy, thoracolumbar region |  |  |
| 720.0 | Ankylosing spondylitis of unspecified sites in spine | M47.817 | Spondylosis without myelopathy or radiculopathy, thoracolumbar region |  |  |
|  |  | M47.896 | Other Spondylosis, lumbar region |  |  |
|  |  | M47.897 | Other Spondylosis, lumbosacral region |  |  |
|  |  | M41 | Scoliosis |  |  |
|  |  | Q67.5 | Congenital Spinal Deformity |  |  |
|  |  | M41.4 | Neuromuscular Scoliosis |  |  |
|  |  | Q76.3 | Congenital Scoliosis |  |  |
|  |  | M40 | Kyphosis and lordosis |  |  |
|  |  | M80.08XA | Age-related osteoporosis with current pathological fracture, vertebra[e], initial encounter for fracture |  |  |
|  |  | M80.08XD | Age-related osteoporosis with current pathological fracture, vertebra[e], subsequent encounter for fracture with routine healing |  |  |
|  |  | M80.08XG | Age-related osteoporosis with current pathological fracture, vertebra[e], subsequent encounter for fracture with delayed healing |  |  |
|  |  | M80.08XK | Age-related osteoporosis with current pathological fracture, vertebra[e], subsequent encounter for fracture with nonunion |  |  |
|  |  | M80.08XP | Age-related osteoporosis with current pathological fracture, vertebra[e], subsequent encounter for fracture with malunion |  |  |
|  |  | M80.08XS | Age-related osteoporosis with current pathological fracture, vertebra[e], sequela |  |  |
|  |  | M48.52XA | Collapsed vertebra, not elsewhere classified, cervical region, initial encounter for fracture |  |  |
|  |  | M48.54XA | Collapsed vertebra, not elsewhere classified, thoracic region, initial encounter for fracture |  |  |
|  |  | M48.56XA | Collapsed vertebra, not elsewhere classified, lumbar region, initial encounter for fracture |  |  |
|  |  | M48.51XA | Collapsed vertebra, not elsewhere classified, occipito-atlanto-axial region, initial encounter for fracture (collapse at the junction of the cervical region with the skull) |  |  |
|  |  | M48.53XA | Collapsed vertebra, not elsewhere classified, cervicothoracic region, initial encounter for fracture (collapse at the junction of the cervical and thoracic region) |  |  |
|  |  | M48.55XA | Collapsed vertebra, not elsewhere classified, thoracolumbar region, initial encounter for fracture (collapse at the junction of the thoracic and lumbar regions) |  |  |
|  |  | M48.57XA | Collapsed vertebra, not elsewhere classified, lumbosacral region, initial encounter for fracture (collapse at the junction of the lumbar and sacral regions) |  |  |
|  |  | M48.4 | Fatigue fracture of vertebra |  |  |
|  |  | S32.0 | Fracture of lumbar vertebra: stress frac.. ***, ** |  |  |
|  |  | S22.0 | Fracture of thoracic vertebra |  |  |
|  |  | S12.0 | Fracture of cervical vertebra |  |  |
|  |  | C41.2 | Primary malignant neoplasm of bone and articular cartilage: vertebrae |  |  |
|  |  | D16.6 | Benign neoplasm of bone and articular cartilage: Spine |  |  |
|  |  | M81.0 | Age-related osteoporosis without current pathological fracture |  |  |
|  |  | M42.1 | Adult osteochondrosis of spine, site unspecified |  |  |
|  |  | A18.01 | Tuberculosis of spine |  |  |
|  |  | M48.2 | Kissing spine, site unspecified |  |  |
|  |  | M43.2 | Fusion of spine, site unspecified |  |  |
|  |  | M48.1 | Ankylosing hyperostosis [Forestier], site unspecified |  |  |
|  |  | M46.5 | Other infective spondylopathies, site unspecified |  |  |
|  |  | M46.9 | Unspecified inflammatory spondylopathy, site unspecified |  |  |
|  |  | M45.9 | Ankylosing spondylitis of unspecified sites in spine |  |  |
